# Telemedicine For Prenatal Care: A Systematic Review

**DOI:** 10.1101/2021.05.14.21257232

**Authors:** Eliza Nguyen, Grace Engle, Shalini Subramanian, Kimberly Fryer

**Affiliations:** University of South Florida College of Medicine; University of South Florida; Department of Obstetrics and Gynecology, University of South Florida College of Medicine

## Abstract

**Objective:** To evaluate the effectiveness of telehealth-enhanced prenatal care.

**Data Sources:** We searched for primary literature in PubMed, EMBASE, and Cochrane Library databases.

**Methods of Study Selection:** Studies were included if they were written in English and used telehealth as an adjunct to or substitute for elements of a comprehensive prenatal care system, with pregnant women as the study population. Studies were excluded if they did not involve comprehensive prenatal care, were not in English, or were abstracts only. Two reviewers independently screened studies by titles, abstracts, and full text. Conflicts were resolved by a third reviewer. Remaining conflicts were resolved by a fourth reviewer. Risk of bias was performed independently by two reviewers. Conflicts were resolved by a third reviewer.

**Tabulation, Integration and Results:** The initial search identified 2707 studies, of which 7 met inclusion criteria. One additional study was identified in the grey literature. Studies included 4 non-randomized controlled studies, 1 randomized controlled trial, 2 qualitative studies, and 1 active clinical trial. Telehealth-enhanced prenatal care included remote monitoring and/or virtual visits. Interventions reduced the number of in-person appointments. Patients and providers had high rates of satisfaction with prenatal care delivered via telehealth. Pregnancy outcomes were similar between the intervention and control groups with the exception of one study identifying higher rates of pre-eclampsia and another showing higher rates of gestational diabetes in the telehealth group. Risk of bias assessment revealed moderate bias in all of the non-randomized studies. The randomized controlled trial had low risk of bias.

**Conclusion:** Telemedicine-enhanced prenatal care may decrease the number of in-person prenatal care visits and increase access to care. Future studies should be done to determine neonatal and maternal outcomes of remote care and to study effectiveness of these interventions for women of color and low socioeconomic status.

## INTRODUCTION

Telemedicine has recently grown in popularity among providers and patients as an alternative to in-person care. A recent systematic review of telehealth interventions in multiple areas of obstetrics and gynecology highlights the expansive nature of technology-assisted care.^1^ While telemedicine encompasses a variety of interventions, including remote monitoring, mobile phone applications, and remote image transmission, the use of virtual healthcare has rapidly expanded due to the COVID-19 pandemic.^2–4^ Given requirements for social distancing and isolation precautions, remote care is favored in situations where it is necessary to minimize exposure, maintain patient volume, and preserve protective equipment. With additional concerns around maternal health in the context of COVID-19, virtual prenatal care has emerged as a priority in obstetrics.^4^ In fact, in August 2020, the American College of Obstetricians and Gynecologists (ACOG) endorsed a bill to remove Medicare restrictions on telehealth and expand coverage beyond the COVID-19 pandemic, suggesting that this type of remote practice will continue to play a role in obstetric and gynecologic care into the future.^5^

Telemedicine has also been recognized as a potential approach to improve disparities in health care among Black and Latinx women, and women in rural communities.^3,6,7^ Telehealth interventions may mitigate barriers to care such as transportation limitations, child care, provider shortages, travel distance, waiting time, and psychosocial stressors.^8,9^ In doing so, these interventions may decrease the overall burden of prenatal care appointments. By providing timely access to care, frequent and convenient engagement, and active patient-focused participation, such interventions can begin to meet the needs of underserved populations.^8^

Current innovations in prenatal care delivery are beginning to incorporate virtual care as a way to augment existing practice. However, information regarding the effectiveness of virtual prenatal care programs is limited. Here we present a systematic review of telehealth-enhanced prenatal care. Though telehealth and telemedicine encompass a multitude of interventions, in this review we focus on comprehensive systems of prenatal care that are augmented by telehealth. This includes interventions involving remote monitoring, virtual visits, and mHealth supported care.

The purpose of this review is to evaluate the evidence on the use of an integrated virtual care model as an alternative method of delivering prenatal care. This review aims to inform providers of obstetric care how this developing technology might be useful to practice. Additionally, we recognize that the findings from this review may be applicable to practitioners of other specialty and primary care areas who supplement obstetric care.

In this review, we address the following question: Is telehealth an effective means of enhancing standard prenatal care?

## SOURCES

Search criteria were based on the study’s inclusion and exclusion criteria, which were developed using the PICOTS (Population, Intervention, Comparator, Outcomes, Time, Settings) framework. A comprehensive literature search was performed to identify primary literature in ClinicalTrials.gov, Cochrane Library, EMBASE, and PubMed. The search strategy, including MeSH terms and keywords, can be found in Appendix A.

## STUDY SELECTION

The Preferred Reporting Items for Systematic Reviews and Meta-Analyses (PRISMA) statement guided the review.^10^ We closely followed the PRISMA method with the exception of not publishing our protocol on a systematic review reporting website. Inclusion and exclusion criteria were determined using the PICOTS framework. Studies were eligible if they reported primary research data (randomized controlled trials, cohort studies, observational studies), included pregnant women, and utilized telehealth to deliver prenatal care. Detailed inclusion and exclusion criteria can be found in Appendix B.

For the purposes of this review, we define prenatal care based on the guidelines developed by ACOG, which states, “A comprehensive antepartum care program involves a coordinated approach to medical care, continuous risk assessment, and psychosocial support that optimally begins before pregnancy and extends throughout the postpartum period.”^11^

Our definition of ‘telehealth’ is based on the ACOG Committee Opinion 798, “Implementing telehealth in practice”, which defines telehealth as, “the technology-enhanced care framework that includes services such as virtual visits, remote patient monitoring, and mobile healthcare.”^4^

All references generated from the initial search were uploaded into an evidence distilling software, Rayyan QCIR.^12^ This software automatically identified duplicate articles and three reviewers, EN, GE, and SS determined whether or not duplicates were correctly identified. Confirmed duplicates were removed.

During the first phase of the review, two reviewers independently reviewed the titles and abstracts of all references based on the predetermined inclusion and exclusion criteria. In the second phase of the review, two independent reviewers evaluated the full text of all remaining papers based on the same inclusion and exclusion criteria. In each phase, conflicts were resolved by the third reviewer. In the event that a decision was unable to be made by the third reviewer, the study was evaluated by a fourth reviewer, KF, and a final determination was made after discussion between all reviewers. Studies were included if they fulfilled all inclusion criteria and did not meet any exclusion criteria. Data from included studies were extracted by two reviewers. EN and GE. Study characteristics included study design, sample size, population characteristics, details of intervention, outcomes, results, and presence of a control group. These data were entered into a summary table. A summary of evidence table was created to highlight study results, limitations, and applicability to the United States healthcare system. We used the United States Preventive Services Task Force Procedure Manual Appendix XII, Summary of Evidence Table for Evidence Reviews to guide our summary of evidence table.^13^

Two reviewers independently assessed the internal validity of selected studies using the Cochrane Collaboration’s tool for assessing the risk of bias for RCTs, and the ROBINS-I tool for assessing risk of bias in nonrandomized studies of interventions.^14,15^ For RCTs, we evaluated the studies based on the following domains: 1) random sequence generation, 2) allocation concealment, 3) blinding of participants and personnel, 4) blinding of outcome assessment, 5) incomplete outcome data, and 6) selective reporting. For nonrandomized studies, we evaluated the following areas: 1) bias due to confounding, 2) bias in selection of participants into the study, 3) bias in classification of interventions for all outcomes, 4) bias due to departures from intended interventions for all outcomes, 5) bias due to missing data for all outcomes, 6) bias in measurement of outcomes, and 7) bias in selection of the reported result. The assessment tool assigned each domain a rating of low, high, or unclear risk of bias. Conflicts regarding risk of bias were resolved by a third reviewer. The risk of bias figure was created using the ROBVIS visualization tool.^16^

## RESULTS

A total of 2707 studies were identified in the initial search. After duplicates were removed, 2067 remained. 2048 were excluded based on title and abstract and 10 were excluded after full text review. One additional study was found in a search of the grey literature (Figure 1). Final included studies are listed in the study characteristics table (Table 1). Our final review includes one RCT, three non-randomized cohort studies, one quality improvement study, two qualitative studies, and one active clinical trial. Six of the studies included were conducted in the United States. The other two were based internationally in Taiwan and Burkina Faso. Summary of findings and evidence is included in the summary of evidence table (Table 2).

**Table 1.**
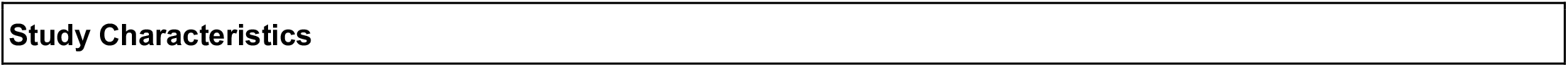
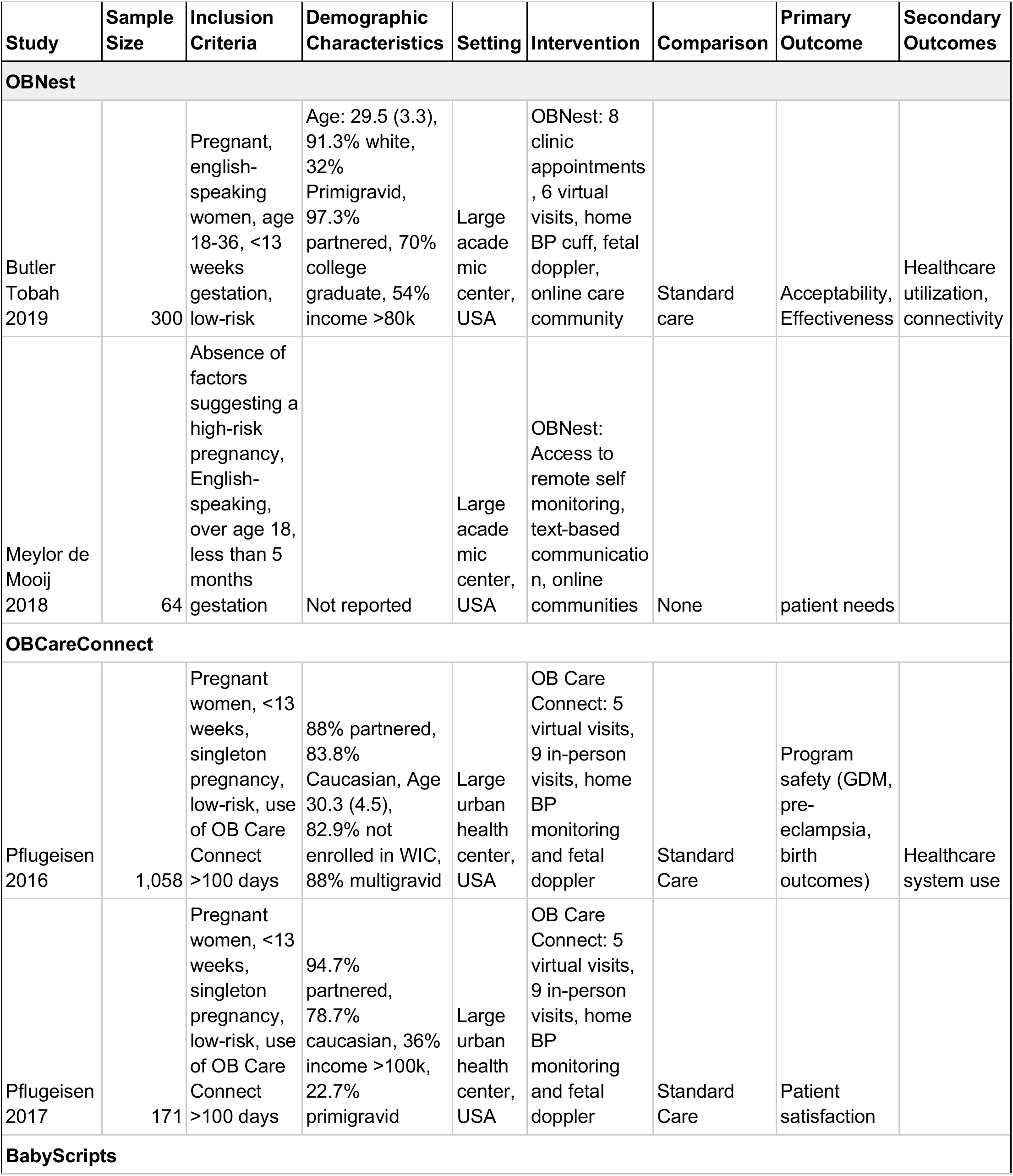
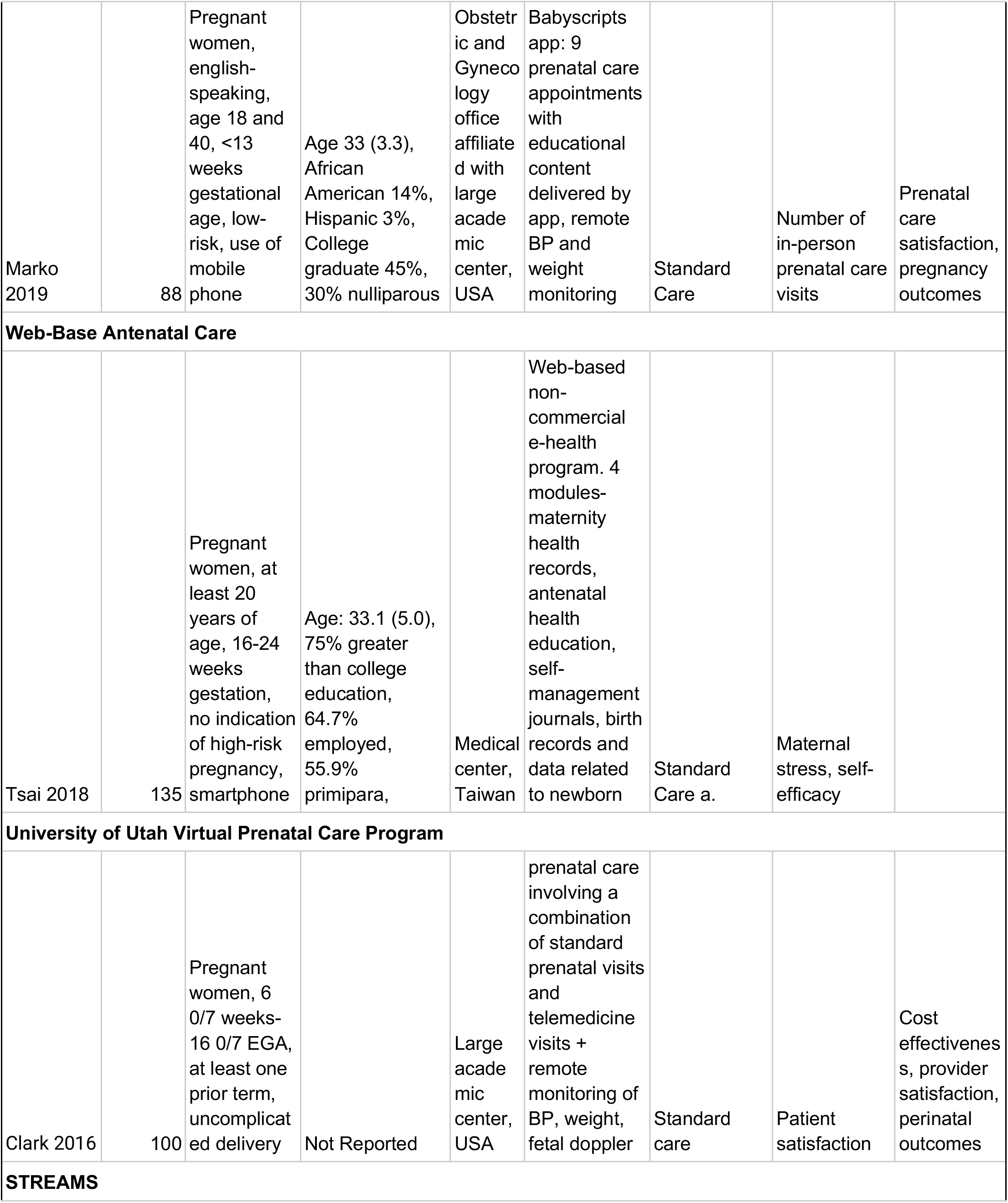
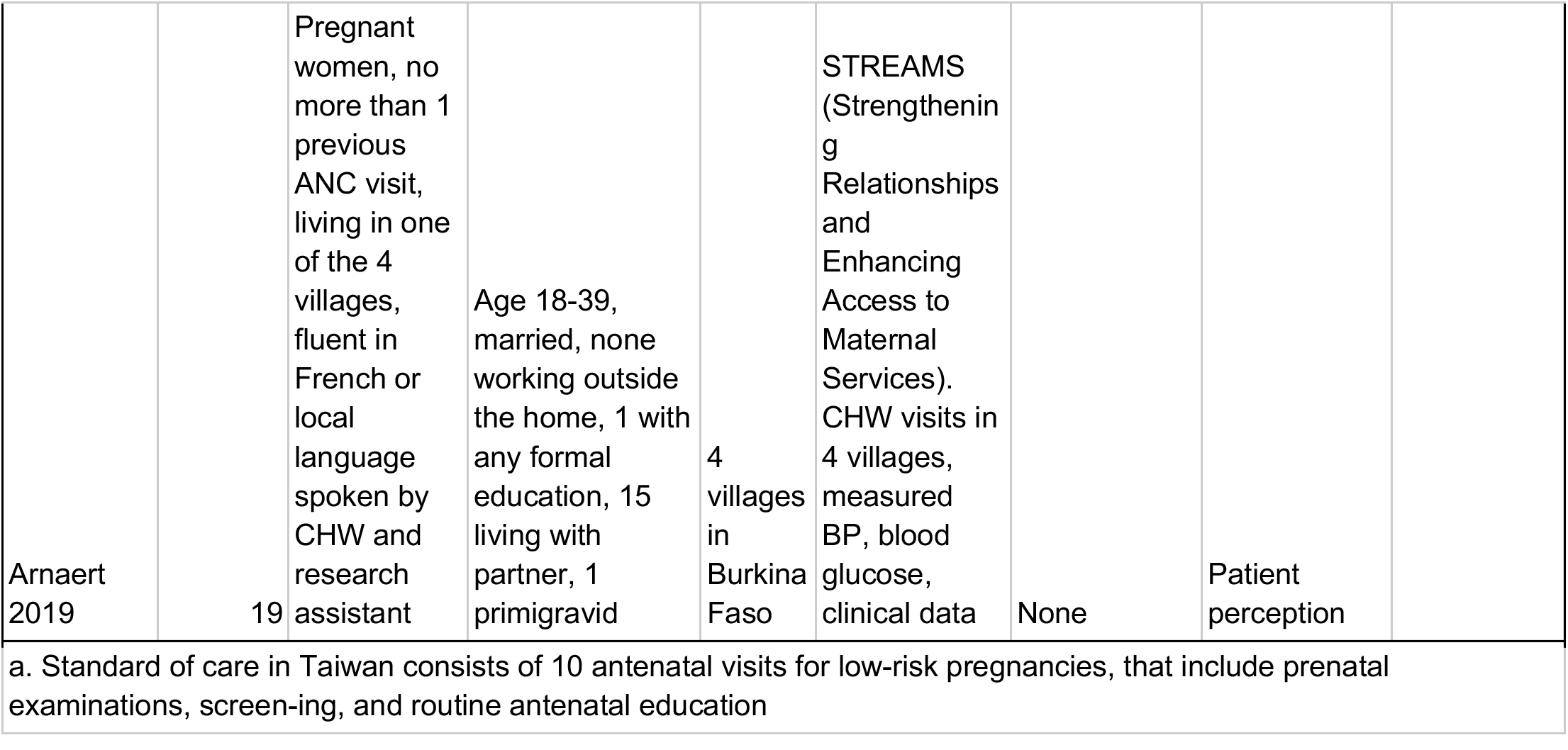
Study Characteristics by intervention.

**Table 2.** Summary of Evidence Table by intervention.

| Summary of Evidence |  |  |  |
| --- | --- | --- | --- |
| Intervention | Summary of Findings | Body of Evidence Limitations | Applicability |
| OBNest |  |  |  |
| Butler Tobah 2019 | Higher satisfaction with care, Lower pregnancy related stress, fewer obstetric provider appointments, higher incidence of GDM in intervention group | Population primarily college-educated, white women, high SES; providers and patients not blinded; underpowered to detect differences in adverse events | Studies conducted in US. Applicable to US healthcare system. |
| de Mooij 2018 | Increased sense of control, confidence, reassurance, high levels of engagement, high satisfaction. | Population primarily college-educated, white women, convenience sampling |  |
| OBCareConnect |  |  |  |
| Pflugeisen 2016 | Higher incidence of pre-eclampsia in intervention group. No difference in birth outcomes, or healthcare system use. | Unequal size of comparison and intervention groups, population primarily caucasian, not WIC enrolled | Studies conducted in US. Applicable to US healthcare system |
| Pflugeisen 2017 | Higher satisfaction in intervention group | Retrospective, voluntary response survey; low response rate (19.8% intervention vs 12.1% control) |  |
| Babyscripts |  |  |  |
| Marko 2019 | No difference in patient satisfaction or provider satisfaction. | Quasi-experimental design, participants allocated based on type of phone, unblinding of participants and investigators, patients mostly college-educated (96% intervention vs 88% control) | Study conducted in US. Applicable to US healthcare system |
| Web-Based Antenatal Care |  |  |  |
| Tsai 2018 | higher self-efficacy, higher satisfaction with antenatal care, higher satisfaction with antenatal guidance in intervention group | Quasi-experimental design, unblinding of participants and investigators, confounders not evaluated, almost all participants married, majority college educated | Study conducted in Taiwan. Moderately applicable to US healthcare system |
| **STREAMS** |  |  |  |
| Anaert 2019 | Intervention beneficial due to early detection of pregnancy-related complications, promoting collaboration between CHWs and midwives, and providing reassurance, peace of mind, and engagement | Small sample size, qualitative study only | Study conducted in Burkina Faso. Less applicable to US healthcare system |

**Figure 1.**
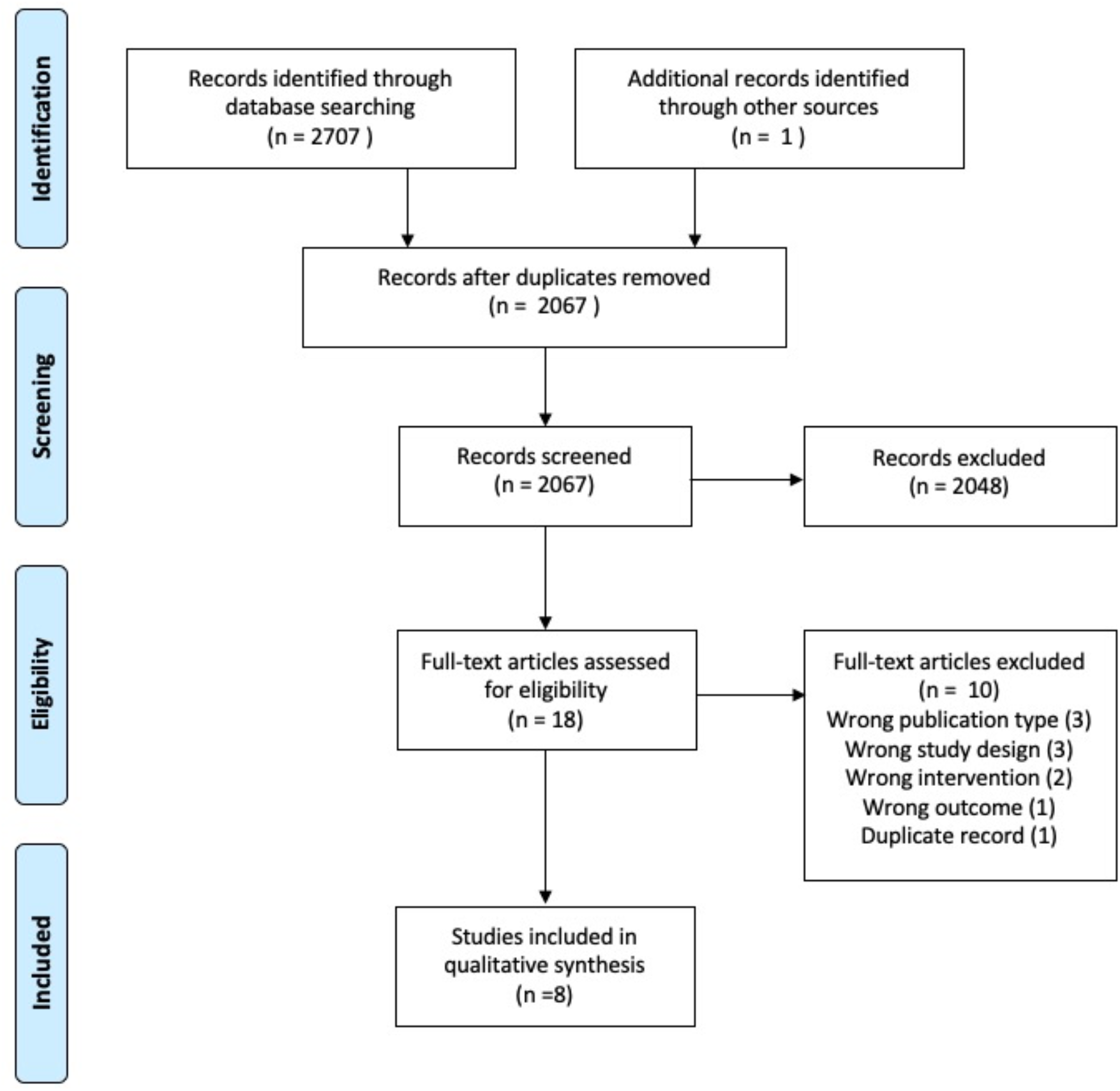
Prisma flow diagram.

**Figure 2.**
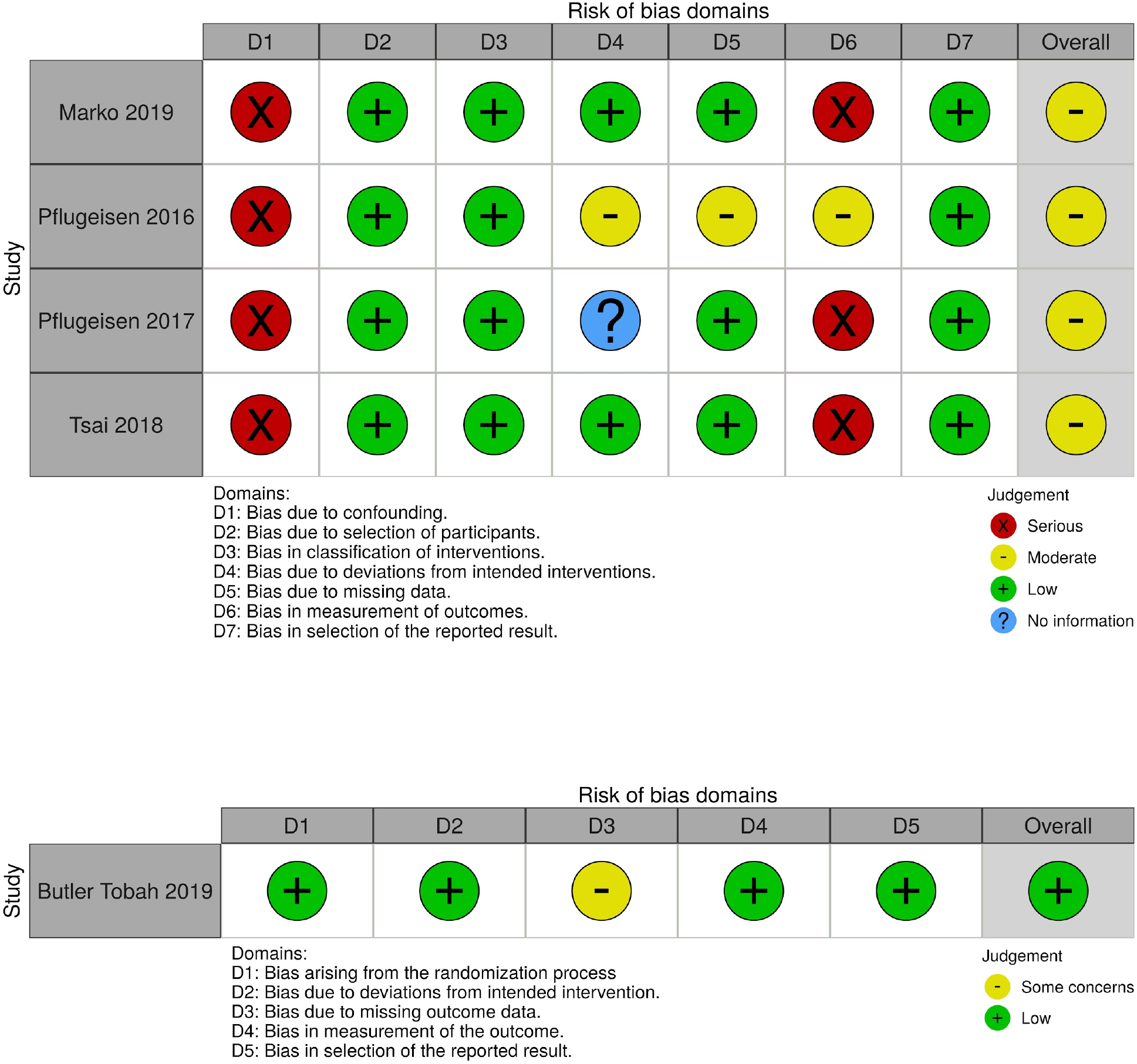
Risk of Bias Assessment by intervention. Includes non-randomized studies (top) and randomized controlled trial (bottom).

### Types of Interventions

Among the papers reviewed, we identified a variety of interventions. Multiple of the studies incorporated smartphone applications that were used for education and remote monitoring.^17–22^ Interventions also largely implemented virtual prenatal visits as an adjunct to in-person care.^17,19–23^ One study used community health workers as an intermediary to report data.^24^ The smartphone applications and remote monitoring systems allowed users to input various health measures such as blood pressure as measured by home cuff, heart rate, body weight, fetal heart rate as measured by home doppler, fetal movements, and uterine contractions. Other interventions used virtual visits in addition to or in place of in person prenatal visits and incorporated a reduced prenatal visit schedule. In the study involving community health workers (CHWs) in remote care, CHWs visited women in rural villages and transmitted data through a remote monitoring system to midwives at the community clinic to monitor for potential complications.^24^

### Prenatal Care Appointment Schedule

Three of the interventions included a reduced schedule of in-person prenatal care appointments.^17–22^ These studies used virtual visits as an adjunct to the traditional in-person care schedule. Two studies implemented a combination of virtual and in-person visits to meet the ACOG recommended number of 12-14 prenatal care appointments.^17–20^ These interventions were able to reduce frequency of in-person care with technology-assisted communication between patients and providers between visits. One study reduced the total number of visits to 8 in-person appointments based on the Department of Defense-Veterans Affairs Uncomplicated Pregnancy Guidelines using remote monitoring to augment care between visits. While the number of prenatal care visits decreased, authors found that more time was spent per visit with higher quality of care for patients receiving prenatal care via telehealth.^21^ Details regarding the exact schedules used can be found in Table 1. The active clinical trial included in this review also implements a combination of virtual and in-person visits.^22^

One study demonstrated that when comparing prenatal care delivered via telehealth with standard prenatal care, there was no difference in ACOG recommended ancillary prenatal care.^17^

### Patient Outcomes

Patient outcomes included acceptability, patient satisfaction, pregnancy-related stress, self-efficacy, effectiveness, number of prenatal visits, safety, and overall experience of the participants.

With regard to acceptability and patient satisfaction, there was either no difference or significantly higher patient satisfaction with the use of the various telehealth interventions.^17,18,21,23^ In one study, 94% of patients reported they would recommend the telehealth program to a friend or relative and 96% said they intended to use the program again. Patients in this study who received the telehealth intervention had increased self-efficacy when compared to patients receiving standard prenatal care.^23^ Another study noted that telehealth participants had lower levels of pregnancy stress.^17^ In the study using community health workers using wireless phones as a liaison between patients and midwives, patients perceived a greater sense of collaboration and felt a sense of reassurance with the use of telemedicine.^24^

Telehealth also proved to be an effective option with regard to patient safety. Multiple studies failed to show significant differences in complications including, cesarean deliveries, preterm delivery, or birth weight between patients receiving standard prenatal care and those receiving telehealth interventions ^17,19,21^ Notably, one study showed a higher rate of preeclampsia in patients participating in virtual visits (8.5% vs 3.4%, p=0.02).^19^ The authors did not comment on potential reasons for or practical significance of the increased rate of preeclampsia in patients enrolled in the virtual track, but they reported no significant difference in other outcomes including mean birthweight, NICU admissions, preterm births, gestational age at birth, or number of cesarean deliveries. They also reported no significant difference in health system use between patients in the virtual and traditional tracks. Of the 10 patients in the virtual track diagnosed with preeclampsia, 8 were diagnosed after 36 weeks and patients were able to remain in the virtual track due to increased proportion in-person physician visits in the third trimester. One study also found an increased incidence of gestational diabetes mellitus (GDM) in the telehealth group as compared to controls.^17^ However, the authors noted that the prevalence of GDM in the intervention group was consistent with that among low-risk pregnancies overall, therefore concluding that the finding represented an artifact that is likely not to be clinically meaningful.

### Provider Outcomes

One study found no difference in provider satisfaction between the intervention and control groups.^21^ Clark et al. are investigating both provider satisfaction and cost-effectiveness in their ongoing clinical trial, the results of which are pending.^22^

### Risk of Bias Assessment

Results of the risk of bias assessment can be found in Figure 3. For non-randomized controlled trials, all were found to have an overall moderate risk of bias. A notable source of bias in these studies included confounding variables that were unaccounted for by the authors. Marko et al. included only iOS users in the intervention arm due to constraints of the mobile application.^21^ Pflugeisen et al did not account for socioeconomic status in their analysis except as determined by WIC status.^19^ They also did not include education level in the analysis. The authors also noted a substantial amount of missing data. The data generated by Pflugeisen and Mou was based on mail-in survey responses, which likely had substantial response bias. Furthermore, response rates in the study were low for both the intervention and the control groups, at 19.8% and 12.1%, respectively.^20^ Finally, Tsai et al did not note any method of controlling for confounding variables.^23^ These studies were also biased in measurement of outcomes. This bias is primarily based on the inability to conceal the intervention from participants or outcome assessors, which could have influenced results.

The RCT by Butler Tobah et al had an overall low risk of bias.^17^ The qualitative studies and active RCT were not included in the risk of bias assessment.

## DISCUSSION

Our review adds to the body of literature highlighting the growing use of telemedicine in obstetrics. We specifically included interventions that incorporate telemedicine into the established prenatal care system, either as an adjunct to or substitute for in-person care. While we identified only a small number of interventions, our findings are promising for future advances in prenatal care.

An important finding from this review is that telemedicine interventions may allow for a reduced schedule of in-person prenatal care visits. Notably, several of the interventions included in this review implemented a schedule with nine or fewer face-to-face prenatal care visits, with virtual visits, remote monitoring, or both, in the intervening time between appointments. This deviates from current ACOG guidelines, which recommend 12-14 prenatal care appointments for women with low-risk pregnancies.^11^ However, other studies have evaluated the safety and efficacy of alternate, reduced prenatal care schedules and have found no associated increase in adverse maternal or perinatal outcomes.^25,26^ A systematic review of a reduced prenatal care schedule revealed lower levels of satisfaction; however our results did not support this finding.^25^ This is likely due to the fact that patients remained connected to their providers remotely between face-to-face visits.

This finding has important implications for delivering prenatal care in unique contexts. Women who are unable to easily access obstetric care may benefit from virtual visits and remote monitoring. A reduced in-person visit schedule augmented by telemedicine has the ability to mitigate the effects of barriers to care, such as transportation constraints, childcare obligations, and work restrictions. These barriers are particularly salient when considering underserved populations such as rural and low socioeconomic status women.^3,8^ Virtual visits and remote data transmission provide flexible options for care that may improve adherence to guidelines. Furthermore, these telemedicine interventions address the rapidly growing need for alternative methods of prenatal care delivery in contexts such as the COVID-19 pandemic where face-to-face visits may be harmful to pregnant women. ACOG has stated that, given the health risks associated COVID-19, modifying the delivery of prenatal care, including through telemedicine, and through a reduced visit schedule, may be appropriate.^27^

The studies included in this review focused largely on patient satisfaction. Results consistently showed either no difference or improved satisfaction with telemedicine interventions when compared to the standard of care. Several studies also identified improvements in self-efficacy and empowerment as sequelae of virtual care. These findings are consistent with literature suggesting that pregnant women enrolled in traditional prenatal care seek information through web-based resources such as mobile apps, blogs, online forums, and search engines as they offer convenient and timely responses in between regular visits.^28,29^ Virtual prenatal care with remote monitoring may offer the resources necessary for women to fill this gap in the prenatal care system. Telemedicine may also serve as a tool to empower women during their pregnancy.

This review has several important limitations. While the studies included here found few adverse events overall, they were not adequately powered to detect significant differences in maternal or neonatal outcomes between the intervention and control groups. This is expected given that the study population included only low-risk pregnancies. Future studies should include a larger sample size in order to assess maternal and neonatal complications in the context of virtual care interventions. This is especially important given the findings of increased rates of pre-eclampsia in one study and increased rates of GDM in another study among virtual care participants. Furthermore, the vast majority of patients in studies conducted in the United States were white women. They generally were of high socioeconomic status and were at least college educated. It is unclear how these interventions would translate to less resourced populations including ethnic and racial minorities and low SES communities. Future studies should include minority and underserved populations in order to better represent the patient population receiving prenatal care. Lastly, one study included in the review was conducted in Burkina Faso, Africa, a lower resourced setting than the United States and Taiwan. It is unclear how that study, which utilizes community health workers, applies to other contexts; however, future research could address this question, particularly in other developing countries.

This review highlights the growing use of telemedicine for the delivery of prenatal care. It specifically assesses telehealth interventions that augment existing prenatal care systems. The studies reviewed here reveal positive outcomes with regard to patient satisfaction and self-efficacy. We anticipate that telemedicine will become an increasingly important part of obstetric care in the future, particularly for low-risk pregnancies. Further studies should be done to include minority and underserved populations.

## Data Availability

No primary data is included

## Appendix A

### PUBMED

(((((((((((((((((pregnancy[MeSH Terms]) OR pregnancy[Title/Abstract]) OR pregnancies[Title/Abstract]) OR gestation[Title/Abstract]) OR pregnant women[MeSH Terms]) OR pregnant woman[Title/Abstract]) OR woman, pregnant[Title/Abstract]) OR women, pregnant[Title/Abstract]) OR prenatal care[MeSH Terms]) OR prenatal care[Title/Abstract]) OR care, prenatal[Title/Abstract]) OR antenatal care[Title/Abstract]) OR care, antenatal[Title/Abstract]) OR perinatal care[MeSH Terms]) OR perinatal care[Title/Abstract]) OR care, perinatal[Title/Abstract])) AND (((((((((((telemedicine[MeSH Terms]) OR telemedicine[Title/Abstract]) OR mobile health[Title/Abstract]) OR health, mobile[Title/Abstract]) OR mhealth[Title/Abstract]) OR telehealth[Title/Abstract]) OR ehealth[Title/Abstract]) OR remote consultation[MeSH Terms]) OR consultation, remote[Title/Abstract]) OR teleconsultation[Title/Abstract]) OR teleconsultations[Title/Abstract])

### EMBASE

‘pregnancy’/exp OR ‘pregnancy’:ab,ti OR ‘child bearing’:ab,ti OR ‘childbearing’:ab,ti OR ‘gestation’:ab,ti OR ‘gravidity’:ab,ti OR ‘intrauterine pregnancy’:ab,ti OR ‘labour presentation’:ab,ti OR ‘labor presentation’:ab,ti OR ‘pregnancy maintenance’:ab,ti OR ‘pregnancy trimesters’:ab,ti OR ‘pregnant woman’/exp OR ‘pregnant woman’:ab,ti OR ‘prenatal care’/exp OR ‘ante natal care’:ab,ti OR ‘antenatal care’:ab,ti OR ‘antenatal control’:ab,ti OR ‘perinatal care’/exp OR ‘care, perinatal’:ab,ti OR ‘perinatal medicine’:ab,ti OR ‘perinatology’:ab,ti AND ‘telemedicine’/exp OR ‘tele medicine’:ab,ti OR ‘telehealth’/exp OR ‘e-health’:ab,ti OR ‘ehealth’:ab,ti OR ‘tele-health’:ab,ti OR ‘teleconsultation’/exp OR ‘remote consultation’:ab,ti OR ‘tele-consultation’:ab,ti OR ‘telephone consultation’:ab,ti OR ‘telemonitoring’/exp OR ‘distant monitoring (patient)’:ab,ti OR ‘distant patient monitoring’:ab,ti OR ‘remote monitoring (patient)’:ab,ti OR ‘remote patient monitoring’:ab,ti OR ‘tele monitoring’:ab,ti

### COCHRANE

(((((((((((((((((pregnancy[MeSH Terms]) OR pregnancy[Title/Abstract]) OR pregnancies[Title/Abstract]) OR gestation[Title/Abstract]) OR pregnant women[MeSH Terms]) OR pregnant woman[Title/Abstract]) OR woman, pregnant[Title/Abstract]) OR women, pregnant[Title/Abstract]) OR prenatal care[MeSH Terms]) OR prenatal care[Title/Abstract]) OR care, prenatal[Title/Abstract]) OR antenatal care[Title/Abstract]) OR care, antenatal[Title/Abstract]) OR perinatal care[MeSH Terms]) OR perinatal care[Title/Abstract]) OR care, perinatal[Title/Abstract])) AND (((((((((((telemedicine[MeSH Terms]) OR telemedicine[Title/Abstract]) OR mobile health[Title/Abstract]) OR health, mobile[Title/Abstract]) OR mhealth[Title/Abstract]) OR telehealth[Title/Abstract]) OR ehealth[Title/Abstract]) OR remote consultation[MeSH Terms]) OR consultation, remote[Title/Abstract]) OR teleconsultation[Title/Abstract]) OR teleconsultations[Title/Abstract])

## Appendix B

Study inclusion criteria are listed below:

Population: pregnant women

Intervention: delivery of comprehensive prenatal care via telemedicine

Comparison: standard of care

Outcomes: Proof of concept, feasibility, effectiveness, provider satisfaction, patient satisfaction, safety, patient health and behavioral outcomes

Types of study design: RCT, clinical trials, cohort study design, qualitative studies

Setting: no restriction

Years of publication: no limit

Publication type: published primary studies, on-going clinical trials

Language: English

Study exclusion criteria are listed below:

Population: non-pregnant women

Intervention: postpartum care, mobile app for management of specific pregnancy-related complications (ex. GDM, gestational hypertension, weight management), interventions involving only educational materials

Types of study design: descriptive studies, systematic reviews, literature reviews Publication type: conference proceedings, abstract only, book chapter review Language: Non-English

